# Intrauterine growth restriction and risk for arterial hypertension in later life. Ribeirao Preto birth cohort study

**DOI:** 10.1101/2024.05.29.24308174

**Authors:** Paulo Cesar Lopes, Paulo Ricardo H. Rocha, Heloisa Bettiol, Marco Antonio Barbieri, Eduardo B. Coelho

**Affiliations:** From the Departments of Internal Medicine (P.C.L., E.B.C) and Pediatrics (P.R.H.R., H.B., M.A.B.), Ribeirão Preto Medical School, University of São Paulo

**Keywords:** intrauterine growth restriction, blood pressure, vascular damage, pulse wave velocity, hypertension

## Abstract

**Background:** Intrauterine Growth Restriction (IUGR) may contribute to the risk of Arterial Hypertension (AH) in adulthood, but its impact after the 3rd decade of life, where environmental factors are prevalent, is still uncertain.

**Methods:** 1,594 individuals with 38 and 39 years-old were evaluated from an original cohort of 6,824 newborns between 1978 and 1979. Biochemical tests, office BP, and anthropometric measurements were done. Also, questionnaires regarding data on income, habits, education, and information about birth were recorded. Krammer’s criteria defined IURG. Subclinical vascular damage was investigated using Pulse Wave Velocity (PWV).

**Results:** The IURG group has higher BP [Systolic BP: 123.3±15.6 vs. 121.2± 13.4 (p=0.049); Diastolic BP: 79.1 ± 10.6 mmHg vs. 76.8 ± 9.9 mmHg (p=0.002)] and higher prevalence of HA [56 vs. 49%; OR=1.30 (1.1-1.8), p=0.04] than the control group. There were no differences in PWV and prevalence of early vascular aging (PWV > 2SD of the mean of normal age value) between groups.

**Conclusion:** IURG was associated with an increase in BP and with a higher risk of hypertension at the end of the third decade of life.

## Introduction

Intrauterine Growth Restriction (IUGR) induces fetal programming due to the necessity of a metabolic economy and can be related to a future risk of arterial hypertension (AH) ^1–8^. A study with mono and dizygotic twins demonstrates that the association between birth weight and AH is independent of genetic and environmental factors at birth and adulthood ^9^. Meta-analysis shows the association between an elevation of the BP or AH in low-weight newborns (<2.500 g)^10,11^ and a rise in BP in newborns with very low-weight (<1.500g) or even on premature <37 weeks when compared to newborns with regular weight ^12^.

Although these and other studies have found a reverse association between birthweight and BP throughout life, this is discreet and may have occurred due to random or selective bias, inadequate adjustment for the current weight, and other confounding factors ^13^. In line, other studies do not show elevation of blood pressure in IUGR ^14,15^. Part of the explanations for those findings relies on the fact that the AH phenotype observed at an adult age is due mainly to environmental factors related to sedentarism and obesity ^16^ and that genetic or present at-birth factors should exert a more negligible impact on the AH occurrence. ^17–20^.

The “COORTES” project refers to a cohort study of birth that began between 1978/79. The current study analyzes the data of its last visit in 2016/2017. Our main goal was to study, at this development stage, where the environmental factors are already established, if the IUGR presence impacts the BP and if it is an independent risk factor for the AH. Also, to evaluate target-organ damage (TOD) ^21,22^, we measured the carotid-femoral pulse wave velocity (PWV) and the estimated Glomerular filtration rate (GFR) in individuals with and without a history of IUGR.

## Methods

### Study population

The cohort was established between June 1978 and May 1979, with the birth registration of 6,827 non-twin alive newborns, which totals 98% of all the births that occurred in Ribeirão Preto, a city located in the countryside of the São Paulo state, the southeast region of Brazil. The last cohort follow-up occurred between 2016/2017, in which 1,954 individuals 38/39 years old were evaluated. More details about this cohort’s characteristics can be obtained from previous publications ^23,24^ The volunteers were evaluated and interviewed by a trained group at the Clinical Research Centre of the Teaching Hospital of the School of Medicine of Ribeirao Preto, University of São Paulo. Standardized questionnaires regarding socioeconomic status were applied. All the participants signed a free informed consent form. The study was approved by the institutional review board (protocol 1.282.710) following the Declaration of Helsinki. The volunteers’ invitation and location process happened in many ways. The primary way was by phone call and sending an invitation letter to the birth address using the contact details collected during the previous cohort evaluations. Furthermore, we looked for potential participants in public and private health system electronic records. We advertised on social media, local newspapers, and television channels, calling people for the follow-up evaluation. All the interested participants belonging to the initial cohort were included.

### Demographic variables and biochemical tests

All the variables in this cohort study were randomly collected or measured, according to the arrival and inclusion on the database, without the research team’s previous knowledge of which group (with IUGR or without IUGR) the volunteers belonged. The BP (mmHg), weight (kg), and height (m) were measured. The body mass index (BMI) was calculated by dividing the weight by height squared (Kg/m²). The waist circumference measurement was obtained by locating the midpoint between the top of the hip bone and the bottom of the rib cage, wrapping a measuring tape around the body at this point, and recording the measurement (in centimeters) with the individual standing upright. Blood samples were collected, processed, and stored in a freezer kept at −80°C until the moment of biochemical tests. Concentrations of serum creatinine, total cholesterol (TC), high-density lipoprotein (HDLc), low-density lipoprotein (LDLc), triglycerides (TG), and glucose were quantified in an automated biochemical analyzer (Weiner, Rosario, Argentina). The Glomerular filtration rate (GFR) was estimated using the serum creatinine values in the CKD-EPI equation. Glycated hemoglobin (HbA1C) was determined by HPLC (Bio-Rad D-10, Hercules, USA). The skin color was auto-inferred and classified as white, black, or brown. Black and brown subjects were analyzed together and classified as non-white. Diabetes mellitus was defined as fasting glucose ≥ 126 mg/dl, HbA1C ≥ 6.5%, or using medication to control the serum glucose. In the same way, Homocysteine > 15 umol/L and C-reactive protein (CRP) > 0.3 mg/dl were considered above the values for normality. The analysis of socioeconomic classes was conducted using the Brazilian Criteria of Economic Classification (BCEC) questionnaire, specifically the 2016 version (Associação brasileira de empresas de pesquisa. Critério Brasil: Critério de Classificação Econômica Brasil 2016: base LSE 2014. http://www.abep.org/criterio-brasil).This questionnaire categorizes individuals into distinct socioeconomic groups based on their household income. For our study, these groups were condensed into four categories, labeled from A to E. Class A represents participants with the highest household income.

The primary data of the initial cohort of 1978/79 were registered using interviews and questionnaires applied to the parents at the moment of birth of the newborn, collecting information related to the mother’s characteristics and state of health, life habits, and socioeconomic condition. Data was collected regarding the number of gestations, maternal educational level (number of years studied), parents’ smoking history, and household income, measured by the family’s salaries standardized by the minimum Brazilian wage salaries per month.

The variable of exposition, IURG, was defined through the Kramer Index, calculated by the ratio between birth weight, measured by a mechanical balance, and estimated weight at the 50 percentile for gestational age, with values below 0,85 defining IURG.

### Outcomes

#### Blood pressure

The BP was measured in 1,594 individuals, of which 233 had IUGR, using calibrated and validated digital sphygmomanometry equipment ^25^ (OMRON 742INT, São Paulo, Brasil). The BP was measured at both arms, with an adequate cuff, 5 minutes after the volunteer had sat down, and the highest of the two values was recorded. The measurement was repeated twice again in five-minute intervals, and the average was calculated.

The prevalence of AH was calculated using the diagnostic criteria recommended by the ACC/AHA 2017^21^. AH was defined when the average between the last two measurements of the SBP was ≥ 130 and/or DBP ≥ 80 mmHg, or when there was previous knowledge of AH, with or without anti-hypertensive medication. To compare BP values, 214 individuals undergoing regular anti-hypertensive therapy were excluded.

#### Pulse Wave Velocity

The measurement of the PWV was made using a portable device (SphygmoCor, AtCor Medical, Sydney, Australia) in all individuals by a trained professional in an environment with controlled temperature and with a rested volunteer on the supine position for at least 5 minutes. The recording of the pulse wave, synchronized by electrocardiographic monitoring, was done using a tonometer at the previously demarcated points in the carotid and femoral areas. The distance between the sternal furcula and the chosen point in the carotid and femoral artery was measured using metric tape, and the software subtracted the distances. The PWV was calculated as the difference between the distance of both measuring spots divided by the temporal lag between 2 pulse waves and was listed in meters per second (m/s).

Besides the measurement of the PWV in both groups, the prevalence of early vascular aging (EVA) was also calculated, taking as a basis two standard deviations beyond the average of the PWV in a healthy population of individuals at the same age, using the value of > 9,2 m/s as a cut-off, based on a European multicenter study.^26^.

### Statistical analysis

The continuous variables results were presented as mean, followed by the respective dispersion measure (standard deviation and 95% of the confidence interval), and the student’s t-test was used to compare groups. Categorical variables were described as percentages (%) followed by the dispersion measure (95% of the confidence interval) and analyzed by chi-square. The exposition effect of IUGR on the BP values was adjusted using a mixed effects generalized linear regression model in three distinct scenarios. The first model incorporated variables with a plausible association with BP rise into the analysis. In this scenario, BMI, body fat percentage, abdominal circumference, sex, skin color, education level, economic category, glycated hemoglobin, GFR, homocysteine, and PWV were employed. In the second model, in addition to the variables previously mentioned, those related to the mother were aggregated, including number of births, smoking during pregnancy, household income, and maternal education level. Only the variables that showed association with IURG in univariate tests were utilized in the third model. The Odds ratio for the same independent factors was calculated using logistic regression analysis. Log Likelihood (LL), Akaike’s (AIC), and Bayesian information criterion (BIC) were used to assess the best fit of the regression models. The P value considered significant was <0.05, and the two-tailed model was chosen. Stata software v.14 (StataCorp, Texas, USA) was used for statistical analysis.

## Results

The main characteristics of the studied sample are summarized in Table 1. Individuals with IUGR were associated, at birth, with maternal smoking history and lower-income households, with a higher proportion of non-white individuals. In adulthood, the economic distribution still predominated in the lower-income groups compared to eutrophic-born individuals. Furthermore, these individuals have higher DBP values (79.1 ± 10.6 mmHg vs. 76.8 ± 9.9 mmHg; p=0.002) and SBP values (123.3 ± 15.6 vs. 121.2 ± 13.4; p=0.049) than the group without IUGR (table 6 suppl.). The exposition effect of IUGR on the BP values was adjusted by mixed effects generalized linear regression model (table 2). The best model was the one that aggregated the maternal variables (tables 3 and 4 suppl.), showing that IUGR exposition, increased abdominal waist, higher pwv, and male gender are associated with higher values of SBP and DBP in adult life. Also, SBP was associated with low household income and low mother’s educational level at birth and with less than 25% of body fat.

**Table 1:** Distribution of the main characteristics from the sample evaluated in individuals with and without IUGR.

|  | No IUGR | IUGR | p-value* |
| --- | --- | --- | --- |
|  | N (%) | N (%) |  |
| *At birth* | 1361 (85.4) | 233 (14.6) |  |
| Mother's educational level |  |  | 0.63 |
| ≥ 8 years | 314 (23.4) | 51 (22.2) |  |
| < 8 years | 1013 (76.3) | 179 (77.8) |  |
| Smoking during pregnancy |  |  | **0.001** |
| No | 970 (74.1) | 133 (63.0) |  |
| Yes | 339 (25.9) | 78 (37.0) |  |
| Number of pregnancies |  |  | 0.69 |
| 1 | 431 (32.5) | 78 (33.9) |  |
| > 1 | 894 (67.5) | 152 (66.1) |  |
| Income in minimum wages |  |  | **0.002** |
| < 6 | 371 (33.4) | 85 (44.2) |  |
| ≥ 6 | 739 (66.6) | 103 (54.8) |  |
| Gender |  |  | 0.49 |
| Male | 664 (48.8) | 108 (46.4) |  |
| Female | 697 (51.2) | 125 (53.6) |  |
| *Current data collection* |  |  |  |
| Race |  |  | **0.04** |
| White | 1083 (79.8) | 171 (74.0) |  |
| Black and mixed race | 274 (20.2) | 60 (7.4) |  |
| Economic classification |  |  | **0.001** |
| A-B | 938 (72.0) | 135 (61.4) |  |
| C-E | 365 (28.0) | 85 (38.6) |  |
| Education level |  |  | 0.81 |
| < 8 years | 964 (76.0) | 168 (76.7) |  |
| ≥ 8 years | 305 (24.0) | 51 (23.3) |  |
| Smoking |  |  | 0.92 |
| No | 1180 (86.8) | 202 (87.1) |  |
| Yes | 179 (13.2) | 30 (12.9) |  |
| Body Mass Index |  |  | 0.24 |
| < 30 kg/m<sup>2</sup> | 872 (64.1) | 158 (68.1) |  |
| ≥ 30 kg/m<sup>2</sup> | 488 (35.9) | 74 (31.9) |  |
| Percentage of Body Fat |  |  | 0.61 |
| < 25% | 1028 (77.9) | 171 (76.3) |  |
| ≥ 25% | 292 (22.1) | 53 (23.7) |  |
| Pulse Wave Velocity |  |  | 0.87 |
| < 9.2 m/s | 1255 (94.9) | 214 (95.1) |  |
| ≥ 9.2 m/s | 68 (5.1) | 11 (4.9) |  |
| Total Cholesterol |  |  | 0.92 |
| < 200 mg/dL | 972 (71.7) | 167 (72.0) |  |
| ≥ 200 mg/dL | 384 (28.3) | 65 (28.0) |  |
| LDLc |  |  | 0.61 |
| < 130 mg/dL | 1046 (80.3) | 175 (78.8) |  |
| ≥ 130 mg/dL | 256 (19.7) | 47 (21.2) |  |
| HDLc |  |  | 0.51 |
| ≥ 40 mg/dL | 901 (68.5) | 157 (70.7) |  |
| < 40 mg/dL | 414 (31.5) | 65 (29.3) |  |
| Blood glucose |  |  | 0.27 |
| < 126 mg/dL | 1269 (93.4) | 212 (91.4) |  |
| ≥ 126 mg/dL | 90 (6.6) | 20 (8.6) |  |
| eGFR |  |  | 0.86 |
| ≥ 90 ml/min/1.73m<sup>2</sup> | 711 (52.0) | 120 (52.0) |  |
| < 90 ml/min/1.73m<sup>2</sup> | 647 (48.0) | 112 (48.0) |  |
| Glycated hemoglobin |  |  | 0.58 |
| < 6.5% | 1312 (96.8) | 223 (96.1) |  |
| ≥ 6.5% | 43 (3.2) | 9 (3.9) |  |
| Homocysteine |  |  | 0.06 |
| < 15 umol/l | 1288 (95.3) | 215 (92.3) |  |
| ≥ 15 umol/l | 64 (4.7) | 18 (7.7) |  |
| C-reactive protein |  |  | 0.22 |
| < 0.3 mg/dl | 852 (62.9) | 136 (58.6) |  |
| ≥ 0.3 mg/dl | 503 (37.1) | 96 (41.4) |  |

**Table 2.** Variables associated with higher Systolic Blood Pressure (SBP) and Diastolic Blood Pressure (DBP) after adjusted by mixed-effects generalized linear regression model. Data express the regression coefficient (CI 95%)

| Variable | SBP |  |  | DBP |  |  |
| --- | --- | --- | --- | --- | --- | --- |
|  | Coef. | CI 95% | p-value | Coef. | CI 95% | p-value |
| IUGR | 2,68 | 0,57 – 4,79 | 0,013 | 2,86 | 1,23 – 4,49 | 0,001 |
| Mother's educational level | -0,43 | -0,83 – -0,03 | 0,036 | - | - | - |
| Household's income | 2,02 | 0,26 – 3,78 | 0,024 | - | - | - |
| Abdominal waist | 0,24 | 0,09 – 0,37 | 0,001 | 0,18 | 0,07 – 0,29 | 0,001 |
| Cf-PWV | 1,42 | 0,88 – 2,00 | <0,0001 | 1,12 | 0,70 – 1,54 | <0,0001 |
| Body fat percentage | -0,18 | -0,32 – -0,03 | 0,018 | - | - | - |
| Male gender | 6,12 | 3,04 – 9,20 | <0,0001 | 3,21 | 0,83 – 5,59 | <0,0001 |

Model adjusted to number of pregnancies, maternal smoking, race, educational level, economic classification, BMI, GFR, Serum homocysteine, Serum C-reactive protein and Glycated hemoglobin.

**Table 3.** Variables associated with risk for hypertension after adjusted by logistic regression model. Values expresse Odds ratios (OR), followed by confidence interval 95% (CI95%)

| **Variable** | **Coef.** | **CI 95%** | **p** |
| --- | --- | --- | --- |
| **IUGR** | 1,66 | 1,12 – 2,45 | 0,011 |
| **BMI** | 2,17 | 1,57 – 3,00 | <0,0001 |
| **Body fat percentage** | 1,99 | 1,36 – 2,91 | <0,0001 |
| **PWV** | 4,74 | 2,05 – 11,0 | <0,0001 |
| **CRP** | 1,75 | 1,28 – 2,40 | <0,0001 |
| **Male Gender** | 3,84 | 2,76 – 5,33 | <0,0001 |

Model adjusted to number of pregnancies, mother's educational level, household's income at birth, abdominal waist, maternal smoking, race, educational level, economic classification, GFR, Serum homocysteine, and Glycated hemoglobin

The prevalence of AH by the criteria of the AHA/ACC was 50.1%. In the univariate analysis, there was an association between the risk of AH and the presence of IUGR exposure [1,33 (95% IC: 1,01-1,76); RR, p=0.044], (table 5 suppl.). A logistic regression model was applied to adjust the variables used in the BP analysis model (table 3). IURG exposure, BMI > 30 m/Kg2, % body fat higher than 25%, pwv higher than 9.2 cm/s, and male gender were associated with risk of AH.

There was not a difference between the PWV of the groups (6,9 ± 1,6 m/s vs. 6,9 ± 1,5 m/s; p = 0.765) or in the prevalence and relative risk to the presence of EVA [5,1% and 4,8%; RR = 1,05 (95% CI: 0,60-1,84), p=0.853], respectively.

## Discussion

The present study shows three main results observed in individuals exposed to IUGR. There was a higher BP when compared to non-exposed individuals. There was an observed higher prevalence of AH in this group, and the presence of IUGR was an independent risk factor for AH.

While the observed office blood pressure difference of 2.0 mm Hg may appear modest, even slight variations in blood pressure hold significance for the population regarding cardiovascular disease prevention^27^. Furthermore, the observed difference in systolic blood pressure (SBP) mirrors those previously documented in individuals born with IUGR during their early years8, indicating that the impact of the intrauterine environment on blood pressure persists into the third decade of life.

In the fourth decade of life, environmental factors, particularly obesity, strongly impact the phenotype of AH. Our population presented a high percentage of overweight/obesity (74%) and AH (50.1%), with a higher prevalence and independent risk for AH in obese individuals [RR = 3.5 (95% CI: 2.8-4.3)], when measured by BMI. Moreover, there was no difference between individuals with and without IUGR regarding BMI, body fat percentage, or obesity prevalence (BMI >30 kg/m2), and these data are consistent with previous publications by other authors.^28,29^. Similarly, male gender, PWV > 9.2 m/s, and C-reactive protein (CRP) > 0.3 mg/dl were independently associated with risk for AH. Of these factors, only CRP was also significantly higher in individuals with IUGR compared to those without IUGR (table 1), and several studies relate higher inflammation measured by CRP as an independent and modifiable marker of cardiovascular risk^30,31,32^.

IUGR was previously associated with an increase in cardiovascular mortality ^33,34,35^. Our cohort’s age between 38 and 39 years old does not allow the prediction of cardiovascular risk through current epidemiological scores since they have not been validated and do not discriminate appropriately for high-risk individuals at this age. Nonetheless, TOD indicates a higher risk of patients being able to receive pharmacological treatment. ^36–40^. The present study uses the PWV measurement to detect subclinical vascular damage. Also, higher PWV values stratified by age have been shown to be an independent risk factor for future cardiovascular events or death ^41^.

Our data demonstrates no difference in the PWV values or EVA prevalence between individuals with and without IUGR. Some non-exclusive hypotheses could explain these findings. One of them is that, at this age, an increase in peripheral vascular resistance due to arteriolar constriction precedes structural and geometric alteration or changes in the elasticity of large vessels, requiring more time for young adults with AH to show significant elevations in the PWV. The second hypothesis would be that the slight BP difference found was insufficient to modify large conductance vessels by the end of the third decade. The assumption that the magnitude of the increase in BP would impact the health of vessels could be inferred by comparing the PWV. In the group of IUGR, Individuals with and without AH have higher PWV (7,1 ± 1,6 vs. 6,6 ± 1,1 m/s; p=0,008), with a difference in the SBP of 21 and DBP of 16 mmHg. Similar results were observed when we replicated this analysis in the group without IUGR or the total cohort.

The choice for the diagnostic criteria of the ACC/AHA (BP ≥ 130/80 mmHg) seemed better than the one adopted by the ESC/ESH (BP ≥ 140/90 mmHg) due to it presenting a higher accuracy in a previous study of our cohort, in the identification of EVA.^42^. Furthermore, the chosen definition could strongly correlate with biochemical risk factors for cardiovascular diseases, arterial stiffness, inflammation, and obesity (tables 1 and 2 suppl.).

This study presents essential qualities. Firstly, by analyzing a representative sample of the original cohort, with no differences in the proportions of the characteristics between the current cohort and the original cohort, such as maternal educational level, smoking during pregnancy, number of gestations, household income, gender, or presence of IUGR. This birth cohort includes 98% of the NB in Ribeirão Preto over one year, making it homogeneous regarding age and eliminating confounding factors. Secondly, due to the reliability of the primary outcome data, extracted from measures obtained with automated digital equipment, validated, and calibrated, performed by trained professionals, and following the AH guidelines recommendations, including rest time and suitability of the cuff to the patient’s arm. In this context, 710 individuals presented BP ≥ 130/80 mmHg, and only 89 individuals were considered hypertensive patients through the questionnaire, reducing the possibility of information bias.

Conversely, the study presents a possibility of systematic errors. One could arise from confounding factors that interfere with the outcome found. Except for family for household income (directly related to the risk of IUGR and AH) and maternal smoking history (risk of IUGR), other possible confounding factors analyzed did not differ between exposition groups when assessed through univariate analysis (table 1). We performed multiple linear and logistic regression models, including household income and maternal smoking, to validate IUGR as an independent association factor in the rise of BP and AH (tables 2 and 3). Nevertheless, we can not rule out possible residual confounding factors (Such as vitamin or protein-calorie deficiency, maternal AH, and diabetes mellitus, among others) that theoretically modify the magnitude of the observed effect.

In conclusion, our study demonstrates that around the fourth decade of life, being born with IUGR is associated with an independent risk for AH and an elevation of the office BP.

## Data Availability

We declare that the data mentioned in the manuscript is open for consultation.

## Notes

### Competing Interest Statement

The authors have declared no competing interest.

### Funding Statement

CNPQ grant number 422504/2016-5 The authors, at no time, received payment or services from third parties for any aspect of the submitted work

### Author Declarations

The volunteers were evaluated and interviewed by a trained group at the Clinical Research Centre of the Teaching Hospital of the School of Medicine of Ribeirao Preto, University of São Paulo. Standardized questionnaires regarding socioeconomic status were applied. All the participants signed a free informed consent form. The study was approved by the institutional review board (protocol 1.282.710) following the Declaration of Helsinki.

